# Safety and Efficacy Concerns of Lopinavir/Ritonavir in COVID-19 Affected Patients: A Retrospective Series

**DOI:** 10.1101/2020.07.23.20153932

**Authors:** Marc-Antoine Lepage, Nicholas Rozza, Richard Kremer, Ami Grunbaum

## Abstract

**Context:** Originally developed for the treatment of human immunodeficiency virus (HIV), the antiviral combination lopinavir/ritonavir (LPV/r) is being investigated for use against coronavirus disease (COVID-19). We present a case series raising safety and efficacy concerns in COVID-19 affected patients.

**Methods:** We measured LPV trough concentrations in 12 patients treated at our center and reviewed their clinical charts for side effects known to occur in HIV patients.

**Results:** Compared to established LPV trough concentrations in HIV treated patients, concentrations in COVID-19 affected patients were 3-fold greater (20.64 +/- 10.14 mcg/mL versus 6.25 mcg/mL). In addition, cholestasis and dyslipidemia toxicity thresholds were exceeded in 12/12 and 11/12 patients respectively. No patients achieved the presumed therapeutic concentration. The side effects noted were mainly gastrointestinal symptoms (5/12, 42%), electrolytes imbalances (4/12, 33%), liver enzyme disturbances (5/12, 42%), and triglyceride elevations (2/12, 17%).

**Conclusion:** None of our patients reached presumed therapeutic LPV concentrations despite experiencing side effects and exceeding cholestasis and dyslipidemia toxicity thresholds. This raises concerns for the safety and efficacy of LPV/r. Clinicians should consider closely monitoring for side effects and not necessarily attribute them to COVID-19 itself.

## Context

Lopinavir/ritonavir (LPV/r) is a human immunodeficiency virus (HIV) protease inhibiting drug combination. It has gained interest for the treatment of severe acute respiratory syndrome coronavirus-2 (SARS-CoV-2) and the associated coronavirus disease (COVID-19). As of July 16, 2020, there were 59 clinical studies of LPV/r in COVID-19 affected individuals posted on the ClinicalTrials.gov website, with accrual goals of over 16,000 patients.

There are 5 adult trials reported in the LPV/r monograph (1), exclusively in the HIV population. Patients were young (mean age 38) and predominantly male (82%).

As of June 20, 2020, our hospital center had hospitalized over 400 patients with COVID-19 with a median age of 74.0 (interquartile range 62.3 to 84.0) and a 48/52 male to female ratio. Of these individuals, 20 were treated with LPV/r. We report our clinical observations and laboratory findings (including measured trough drug levels) in a subset of these individuals.

## Objectives

1. To monitor drug levels and potentially associated side effects in COVID-19 patients hospitalized and treated with LPV/r at our institution.
2. To extrapolate on the efficacy of LPV/r in this population based on measured drug concentrations.

## Methods

In the context of a clinical trial (clinicaltrials.gov/ct2/show/NCT04330690), COVID-19 patients were offered treatment with LPV/r 400/100 mg twice daily (as per HIV treatment regimen) for 14 days or until hospital discharge.

Due to an unexpected complication in a COVID-19 patient receiving LPV/r, a series (n=12) of SARS-CoV-2 infected patients treated with LPV/r were investigated to determine LPV troughs and clinical and laboratory abnormalities possibly related to this therapy.

Plasma trough measurements were performed with a clinically validated liquid chromatography - tandem mass spectrometry (LC-MS/MS) assay. Charts were reviewed for side effects known to occur in HIV patients (1) with an onset of at least 24 hours after initiation of treatment.

## Findings

Gastrointestinal symptoms were observed in 5/12 patients (42%), electrolyte, liver, and triglyceride disturbances in 4/12 (33%), 5/12 (42%), and 2/12 patients (17%) respectively.

Trough concentrations (12 hours post dosing) were measured 4 to 7 days after treatment initiation in 10 of 12 patients. Patient 10 had a trough measurement on day 2, prior to discharge and cessation of treatment. Patient 6 received 12,000 mg of LPV in a single treatment dose due to an error in dosing on day four of treatment (LPV concentrations 34 hours post overdose in this patient remained elevated at 34.5 mcg/mL). Mean LPV trough concentrations were higher than the expected concentrations in HIV patients (20.64 mcg/mL [standard deviation 10.14] vs 6.25 mcg/mL). All 12 COVID-19 patients exceeded reported cholestasis toxicity thresholds (6.43 mcg/mL) (2) and 11/12 (92%) exceeded reported dyslipidemia toxicity thresholds (9.71 mcg/mL) (3).

## Discussion

Schoergenhofer has previously reported that COVID-19 patients treated with LPV/r have higher LPV trough concentrations than HIV patients treated with this antiviral drug combination (4). Furthermore, nearly all trough concentrations in our patients exceeded the toxicity thresholds for cholestasis (2) and dyslipidemia (3). High concentrations may also increase the frequency and severity of other potential complications in these already ill patients.

LPV is 98-99% protein bound. In the absence of human serum (i.e. 100% freely available drug), the in vitro half-maximal effective concentration (EC50) for LPV against SARS-CoV-2 is 16.74 mcg/mL (5) compared to 0.006 - 0.017 mcg/mL for various strains of HIV (1) which is several hundred fold higher. This concentration is unlikely to be obtained in vivo without excessive risk of toxicity, as an accidental overdose of 30 times the intended amount resulted in presumably sub-therapeutic levels. This was despite development of significant toxicities, including acalculous cholecystitis and hypertriglyceridemia.

## Conclusion

Our report supports previous findings that treating COVID-19 affected patients with standard LPV/r dosages (as per routine use in the HIV population) results in higher trough concentrations which may cause more adverse events. Despite this potential risk for drug-related toxicity, the in vitro derived EC50 for LPV on SARS-CoV-2 was not reached in our patients. Supporting these findings, on July 4, 2020, the World Health Organization reported the Solidarity Trial’s recommendation to immediately discontinue the LPV/r treatment arms due to futility and safety concerns (6).

Most importantly, this report will have an immediate impact on patients enrolled in other trials or who may be offered this treatment as part of their standard clinical care. Our findings should alert clinicians to pay closer attention to possible LPV/r side effects and complications in this population. This will enable early detection of toxicity and allow for therapeutic adjustment and possible discontinuation of LPV/r. Consequently, we feel that the timeliness of this report is critical.

## Data Availability

Data referred to in this article has been anonymized and is kept with the study investigator.

